# Sodium intake and high blood pressure among adults on caloric restriction: a multi-year cross-sectional analysis of the U.S. Population, 2007-2018

**DOI:** 10.1101/2020.12.27.20248919

**Authors:** Jorge Andrés Delgado-Ron, Patricio López-Jaramillo, Mohammad Ehsanul Karim

## Abstract

**Aim:** Small studies have shown reduced sodium sensitivity of blood pressure in obese adolescents on caloric restriction. However, no study at the population level has studied such an effect. We aimed to explore the association between mean daily sodium intake and prevalent hypertension among a nationally representative sample of U.S. adults on caloric restriction who participated in the National Health Examination and Nutrition Survey over the last twelve years.

**Methods and Results:** We used a design-based regression model to explore the association between sodium intake and prevalent hypertension. We also conducted sensitivity analyses using multiple imputation chained equations and propensity score matching. We also measured the effect of a binary exposure derived from two widely recommended thresholds of sodium intake: 2.3 and 5.0 grams per day. Among 5,756 individuals, we did not detect any significant association between increased sodium and the odds of hypertension (OR: 0.97; CI 95%: 0.90; 1.05). All our sensitivity analyses are consistent with our main findings.

**Conclusion:** Our findings suggest that people on caloric restriction—a component of healthy weight loss—would see no benefit in reducing sodium intake to lower blood pressure. These results highlight the need to explore new population-specific strategies for sodium intake reduction, including new dietary prescription approaches to improve dietary adherence and reduce the risk associated with sodium-deficient diets.

## Introduction

Hypertension causes more death and disability than any other risk factor globally, ahead of smoking, high glucose, and obesity. Nearly 10.4 million deaths worldwide were attributable to elevated blood pressure in 2017.^1^ Ambard and Beaujard theorized a potential association between salt consumption and hypertension as early as 1904. However, contradictory evidence from observational studies using low-salt diets led to an intense debate that lasted more than a century.^2^ Only recently, well-designed clinical trials provided reliable answers. The Dietary Approaches to Stop Hypertension-Sodium (DASH-Na) trial concluded that “blood pressure can be lowered … by reducing the sodium intake.”^3^ Long-term cohort studies support the DASH-Na trial findings.^4^

The precise mechanisms that cause dietary sodium to modulate blood pressure levels are not well defined, which often translates to uncertainty about who would benefit from such interventions. Past studies have shown the role of specific modifiers like ethnicity, even at the molecular level.^5^ However, we know much less about the role of energy balance—the equilibrium between calories consumed and calories burned through physical activity^6^—as a modifier of “salt sensitivity” of blood pressure.

Energy balance is arguably a more critical modifier than diet or exercise on their own. After all, we reach metabolic tipping points that enable our bodies to regulate blood pressure through the additive interaction of energy intake and energy expenditure.^7,8^ Current studies lack this holistic understanding and, as a result, present somehow conflictive evidence. For instance, a reanalysis of the DASH-Na data found less energy intake associated with increased salt sensitivity.^9^ Similarly, a community-based study in China found that participants in the highest quartile of physical activity had reduced salt sensitivity compared to the lowest quartile. However, there was not a linear association across quartiles.^10^

Caloric restriction and fitness are recommended to prevent or control hypertension.^11,12^ The current European and American hypertension guidelines recommend a reduced sodium intake on top of that.^13,14^ However, it is not clear if patients would benefit from reducing sodium in their diet once they are on an energy deficit. Caloric restriction—a negative energy balance—reduces blood pressure levels through improved insulin sensitivity, reduced adiposity, and reduced sympathetic activity, independent of reaching an ideal body weight.^15,16^ These mechanisms might also play a role in modulating the salt sensitivity of blood pressure.^12,17^ Studies that induced weight loss have modified the relationship between sodium and blood pressure. However, they used small samples and are outdated.^16,18^

In the present work, we used a design-based regression model to explore the association between mean daily sodium intake and prevalent hypertension among U.S. adults on caloric restriction in the National Health Examination and Nutrition Survey (NHANES) from 2007 to 2018.

## Methods

### Data source, Design, and Study Population

We used publicly available data from NHANES, a cross-sectional four-stage stratified cluster complex survey, representative of the non-institutionalized United States population. NHANES gathers lifestyle and medical information, along with biological samples and a physical examination. Readers can find relevant description design and sampling procedures associated with this survey elsewhere.^19^ The study complies with the Declaration of Helsinki and it is covered by item 7.10.3 in the University of British Columbia’s Policy 89 on studies involving human participants and Article 2.2 in the Tri-Council Policy Statement Ethical Conduct for Research Involving Humans.

### Analytic sample and study variables

We included adults aged 20-79 years from six two-year cycles (2007-2018) with an energy deficit of at least -350 calories per day. We chose this cut-off to correct potential underreporting from self-reported dietary data.^20^ We derived energy balance from its two essential components: energy input (mean daily intake) and energy output (basal metabolic rate plus physical activity).^6^ The basal metabolic rate was calculated using the revised Harris-Benedict equations.^21^ Self-reported weekly vigorous and moderate physical activity was transformed into daily metabolic equivalents following NHANES’ suggested scores.^22^ NHANES derives the total daily intake from two 24-hour recall interviews, 3 to 10 days apart, using a validated instrument to reduce recall bias.^23^ We did not exclude participants with invalid or missing answers for physical activity; instead, we used their basal metabolic rate as total output.

We excluded pregnant women and participants with body mass index (BMI) below 18.5 (malnutrition) or an active thyroid pathology due to inherent metabolic differences in these patients. Adults over the age of 80 were excluded because their age is not publicly available due to privacy concerns. We also excluded people who reported a “much less than usual” intake during either interview.

The primary outcome was hypertension, a binary variable indicating one of the following: self-reported use of antihypertensive medication or systolic hypertension (mean systolic blood pressure ≥ 130 mmHg) or diastolic hypertension (diastolic blood pressure ≥ 80 mmHg).^11^ Individuals without a second valid measurement for either systolic (SBP) or diastolic blood pressure (DBP) were excluded to avoid measurement error.

The exposure was self-reported mean consumption of sodium (grams per day) as a continuous measure. Two additional binary variables were derived for our sensitivity analyses (described later). Other covariates included demographic data (age, gender, education, income, and ethnicity), pre-existing diseases (diabetes and obesity), and smoking.^24^ We excluded “Refused,” “Don’t know,” and “Missing” values for demographic variables. However, for diabetes, we required respondents to provide a definitive “Yes.” The details of the analytic data creation process are shown in Figure 1.

**Figure 1.**
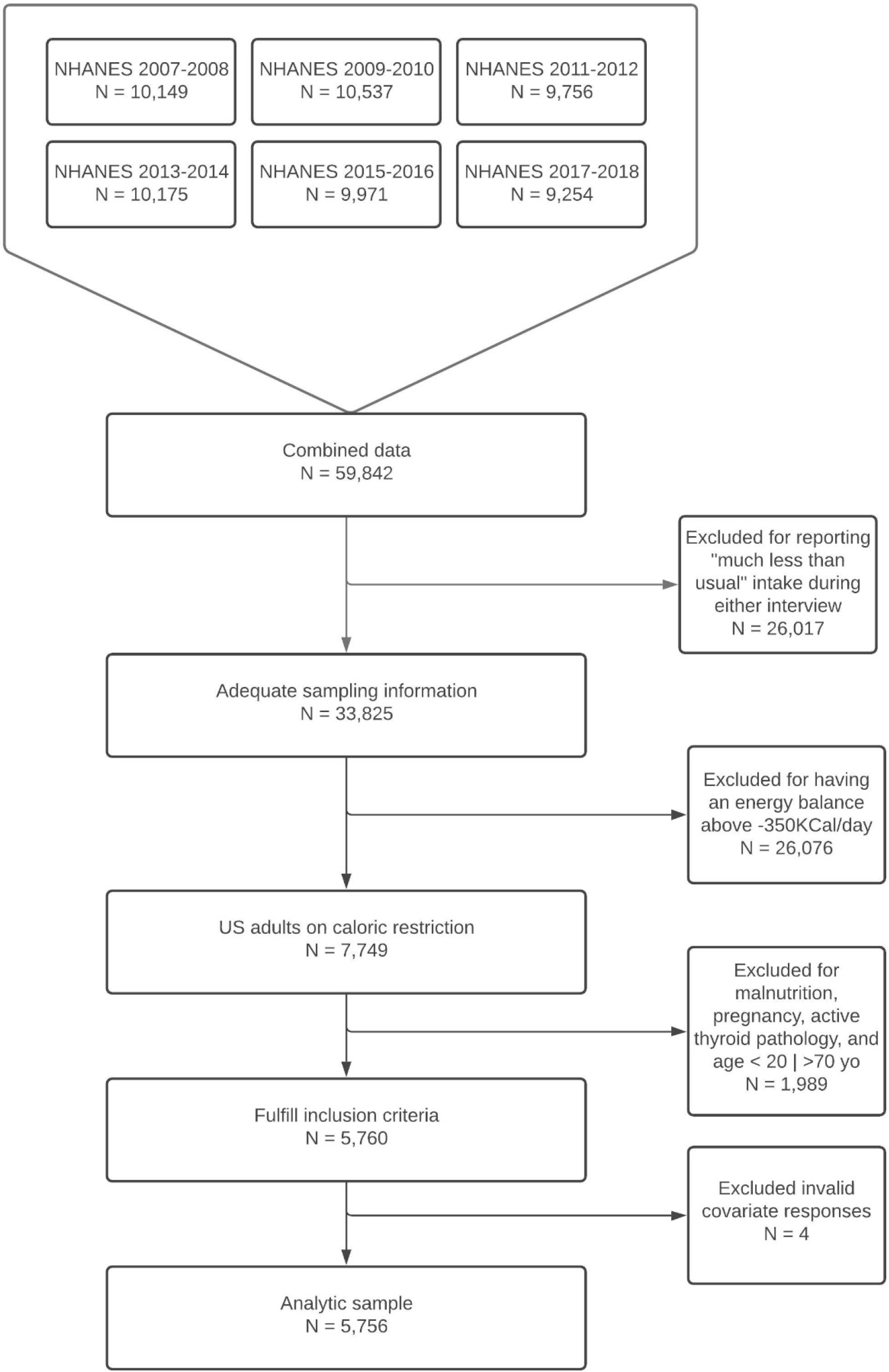
Flow chart of the analytic sample selection for testing the association between sodium intake and high blood pressure among U.S. adults on caloric restriction aged 20-79, using data from the National Health and Nutrition Examination Survey; cycles 2007-2008 to 2017-2018.

Figure 2 depicts our hypothesized causal diagram describing the causal relationships among the variables under consideration. Given we already restricted the sample by energy balance, we minimized bias adjusting for age, diabetes, education, ethnicity, gender, energy expenditure, BMI, and smoking status based solely on Pearls’ backdoor criterion.^25^ Serum sodium and the renin-angiotensin-aldosterone system (unmeasured) were assumed mediators. Other unmeasured variables include alcohol consumption and fat accumulation.

**Figure 2.**
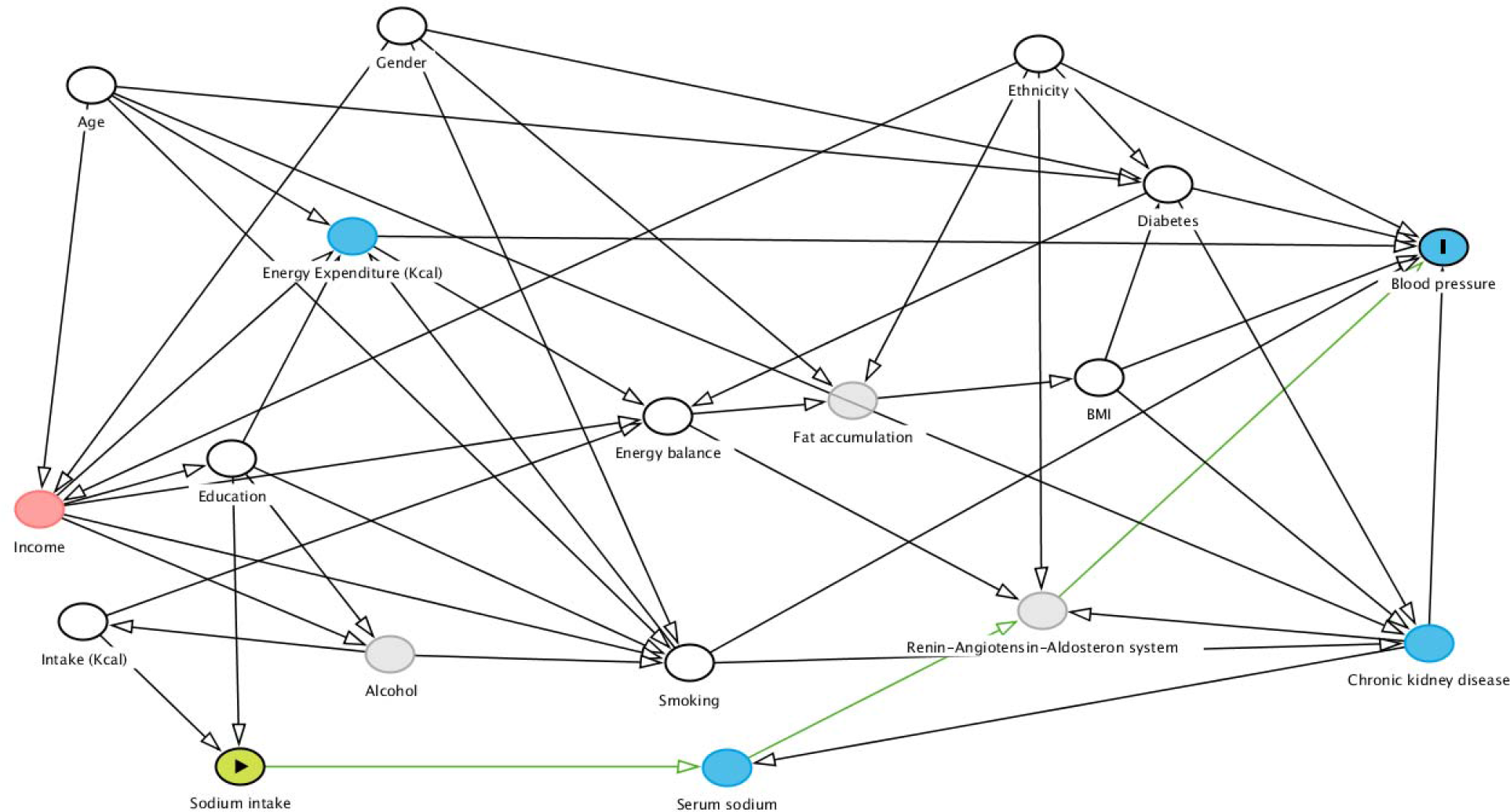
Directed acyclic graph for assessing the causal association between sodium intake and elevated blood pressure. Considering our energy balance restriction, age, gender, ethnicity, education, income, and total intake act as confounders. We achieved sufficient adjustment by controlling age, diabetes, education, ethnicity, gender, energy expenditure, body mass index, and smoking status. Fat accumulation, alcohol, and the renin-angiotensin-aldosterone system (RAAS) are unmeasured variables. Serum sodium and RAAS are mediators. The figure was drawn using the web-version of DAGitty.

### Primary Analysis

We compared individuals’ characteristics by hypertension status (yes/no) using survey-featured *t-*tests for continuous variables and the Rao-Scott F-adjusted χ2 test for categorical variables.^26^ We combined the survey weights from our six cross-sectional subsamples following NHANES recommendations^19^ and built our design using all survey features, subsetting only the eligible sample. We estimated the Odds Ratio (OR) and respective confidence intervals (CI 95%) using a design-based multivariate logistic regression (hereafter referred to as the ‘outcome model’), adjusting for the variables selected based on Pearls’ backdoor criterion as described above.

We probed interactions between sodium intake and the other covariates using Bauer’s inferential and graphical techniques^27^ based on findings from the previous literature.^9^ Collinearity was assessed via variance inflation factors (VIF) and classified following the recommendations by Belsley.^28^

The goodness of fit for all models was evaluated using weighted ROC curves and the Archer-Lemeshow test.^29,30^ All 95% confidence intervals (CI) and Cox-Snell pseudo-R^2^ account for both the survey design and day-to-day dietary intake variations.^19,31,32^

### Sensitivity Analyses

#### Missing data

As missing completely at random assumption may not be realistic, we applied the multivariate imputation by chained equation method.^33,34^ We imputed 20 datasets under the missing at random assumption using all the variables from our primary analysis model as predictors for the imputation model. Rubin’s rules were used to pool the estimates together.^34^

#### Binary exposure based on recommended thresholds

We transformed the exposure in our outcome model to a binary variable. First, we used the threshold of ≥ 2.3 g/day to classify exposure as “high” or “low,” following the maximum sodium intake recommendations in current European and American hypertension guidelines.^13,14^ Then, we used a threshold of ≥ 5.0 g/day, the daily sodium intake recommended by the World Health Organization,^35^ using the same specifications described below.

We performed the analysis with both the complete-case dataset and the imputed datasets, pooling the estimates as described above. We also matched the probability of being exposed to “high” or “low” sodium intake using propensity scores for both thresholds. We performed a 1:1 nearest neighbour match (without replacement) on the propensity score’s logit with a 0.2 calliper.^36^ We modelled the exposure using our original covariates—cycle and survey features were also covariates following Dugoff’s method^37^—to achieve adequate standardized mean difference balance (SMD < 0.2). The survey-featured outcome model was used to estimate the population average treatment effect from the matched subsample. Again, the exposure was binary (“high” vs. “low”).

All statistical analyses were performed using R 4.0.3;^38^ the code is available on request.

## Results

### Study sample characteristics

We evaluated a total of 5,756 individuals, representing a U.S. national population of 53,036,129. The weighted prevalence of hypertension in our sample was 42.6%. On average, hypertensive individuals were significantly more likely to be male, older, diabetic, and overweight or obese. Hypertensive and non-hypertensive people also differed significantly in ethnicity and smoking history, but not education or daily total energy expenditure. Sodium intake did not significantly differ by hypertension status (Table 1).

**Table 1.** Participants characteristics by hypertensive status, U.S. adults who reported caloric restriction aged 20-79. National Health and Nutrition Examination Survey (NHANES) 2007-2018.

|  | Non-hypertensive<br>(n=3,023) | Hypertensive<br>(n=2,733) | *P*-value |
| --- | --- | --- | --- |
| Sodium intake (g/day) [mean (SD)] | 3.20 (1.35) | 3.17 (1.36) | 0.462 |
| Age in years [mean (SD)] | 38.32 (13.57) | 51.28 (14.69) | <0.001 |
| Female (%) | 1874 (47.0) | 1352 (35.5) | <0.001 |
| Ethnicity (%) |  |  | <0.001 |
| White | 1619 (64.3) | 1416 (65.7) |  |
| Latino | 1107 (17.4) | 735 (11.9) |  |
| Black | 830 (12.2) | 1120 (16.3) |  |
| Other | 382 (6.2) | 268 (6.1) |  |
| Education (%) |  |  | 0.12 |
| Some school | 812 (13.7) | 902 (16.8) |  |
| High school degree | 946 (24.3) | 940 (26.5) |  |
| Higher education | 2180 (62.0) | 1697 (56.7) |  |
| Body mass index (kg/m<sup>2</sup>) | 29.56 (6.65) | 32.41 (7.35) | <0.001 |
| [mean (SD)] |  |  | <0.001 |
| Normal | 1008 (27.5) | 397 (12.6) |  |
| Overweight | 1263 (32.3) | 1029 (30.0) |  |
| Obesity Class 1 | 914 (23.0) | 987 (28.2) |  |
| Obesity Class 2 | 435 (9.9) | 560 (14.4) |  |
| Obesity Class 3 | 318 (7.3) | 566 (14.7) |  |
| Diabetes (%) | 234 (3.8) | 827 (18.1) | <0.001 |
| Smoking (%) |  |  | <0.001 |
| Never smoker | 2287 (59.1) | 1813 (51.2) |  |
| Past smoker | 680 (18.1) | 1030 (28.4) |  |
| Current smoker | 971 (22.7) | 696 (20.4) |  |
| Daily energy expenditure (Kcal) | 3450.90 (1867.56) | 3381.06 (1809.72) | 0.314 |

Sample sizes (n) are unweighted. Means and prevalences are weighted but not adjusted by age.
NHANES 2007-2018 participants aged 20-79 years old on caloric restriction (-350Kcal/day) were included; people with active thyroid pathology, malnutrition, and pregnant women were excluded.
Hypertension was defined as mean systolic blood pressure $\geq 130$ mmHg, mean diastolic blood pressure $\geq 80$ mmHg, or self-reported use of hypertensive medication. Two-tailed *P*-values derive from weighted t-tests for continuous variables and the Rao-Scott F-adjusted $\chi^2$ test for categorical variables.

### Association between mean daily sodium intake and prevalent hypertension

A one-gram increase in daily sodium intake did not significantly correlate with higher odds of hypertension in the U.S. population on caloric restriction. The estimate was imprecise around the null in both the crude (OR=0.98, 95% CI: 0.93; 1.03) and adjusted models (Table 2). We did not find any significant interaction between the exposure and other covariates. Multicollinearity was “near weak” for all independent variables (VIF < 2.5).

**Table 2.**
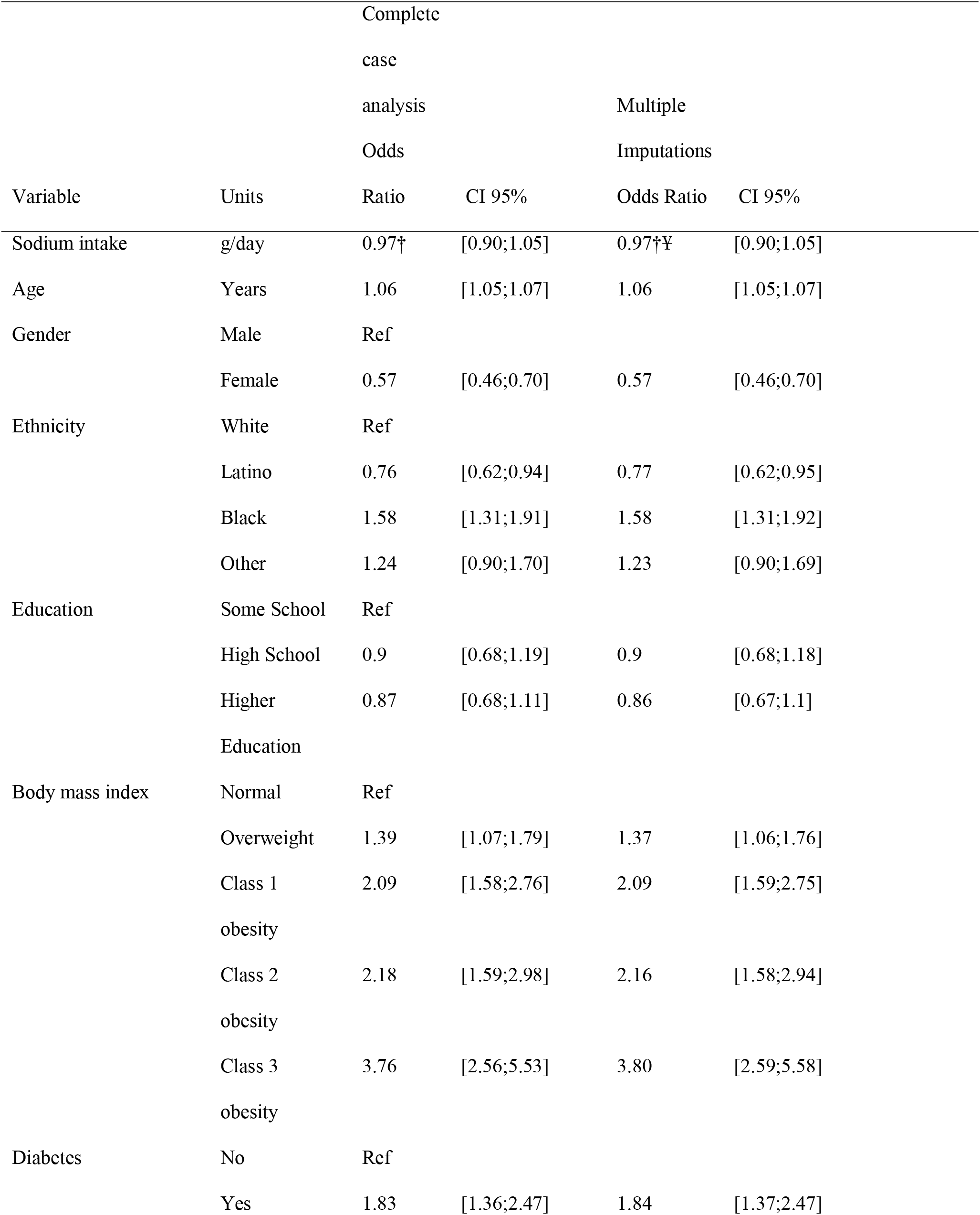
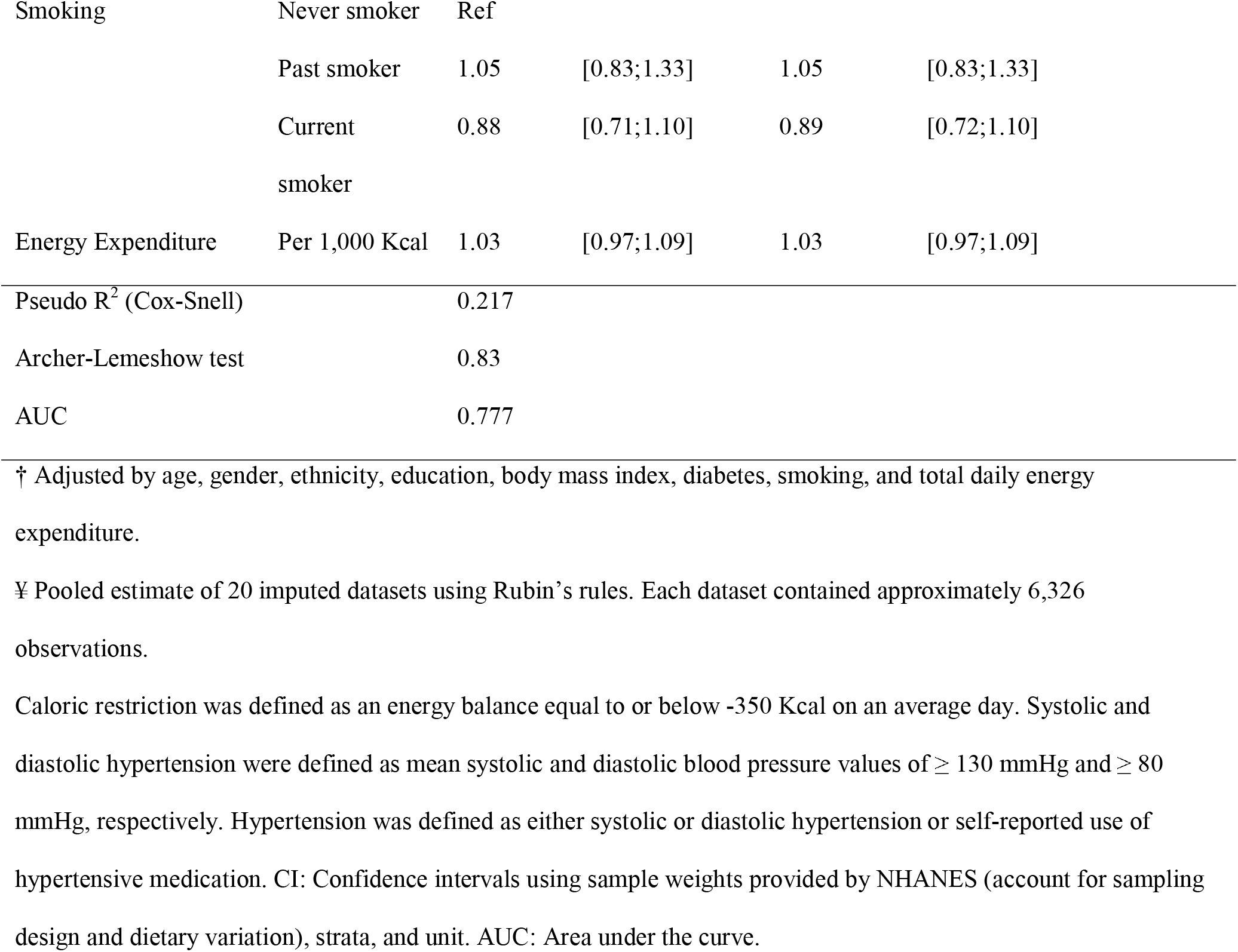
Survey-featured multivariable logistic regression model for the relationship between average daily sodium intake (grams per day) and hypertension among U.S. adults in caloric restriction, aged 20-79. National Health and Nutrition Examination Survey (NHANES) 2007-2018

### Sensitivity analysis

#### Multiple imputations for missing data

Data for moderate and vigorous physical activity was missing for 30% of the sample. Other sampling variables with lower proportions of missing data were calorie intake (9%), height (8%), and the derived BMI (8%). Model variables with missingness were mean sodium intake (9%) and education (40.8%). Each imputed dataset contained approximately 6,326 observations. The estimate and confidence intervals were equal to that of the primary analysis (OR=0.97, 95% CI: 0.90; 1.05) (Table 2).

#### Sodium intake of ≥ 2.3 g/day

Consuming 2.3 grams of sodium per day or more was not significantly associated with higher hypertension odds in either the complete-case or imputed data analyses. Although both estimates found a different effect, they were imprecise around the significance threshold. Hence, we failed to find an association (Table 3).

**Table 3.**
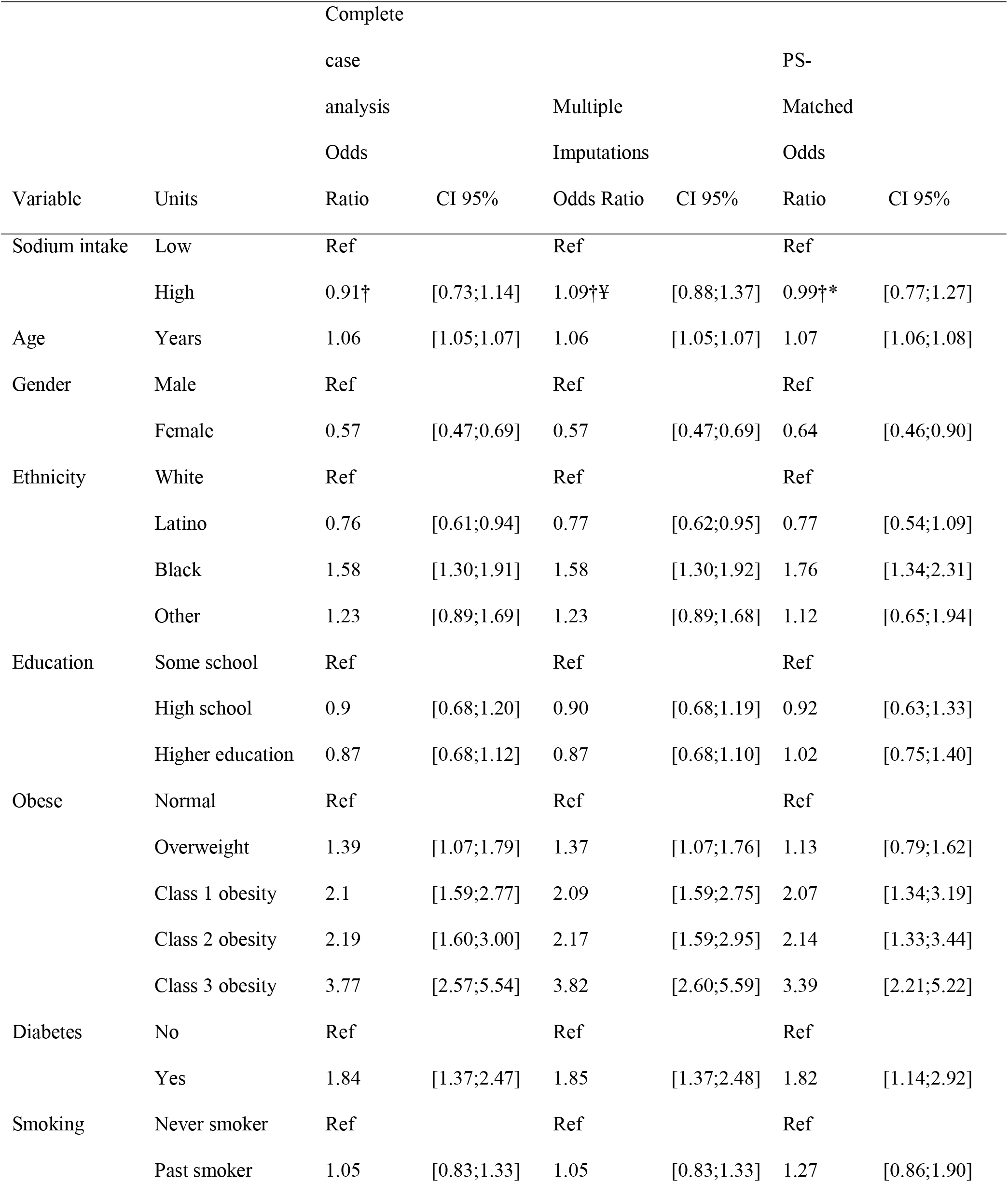
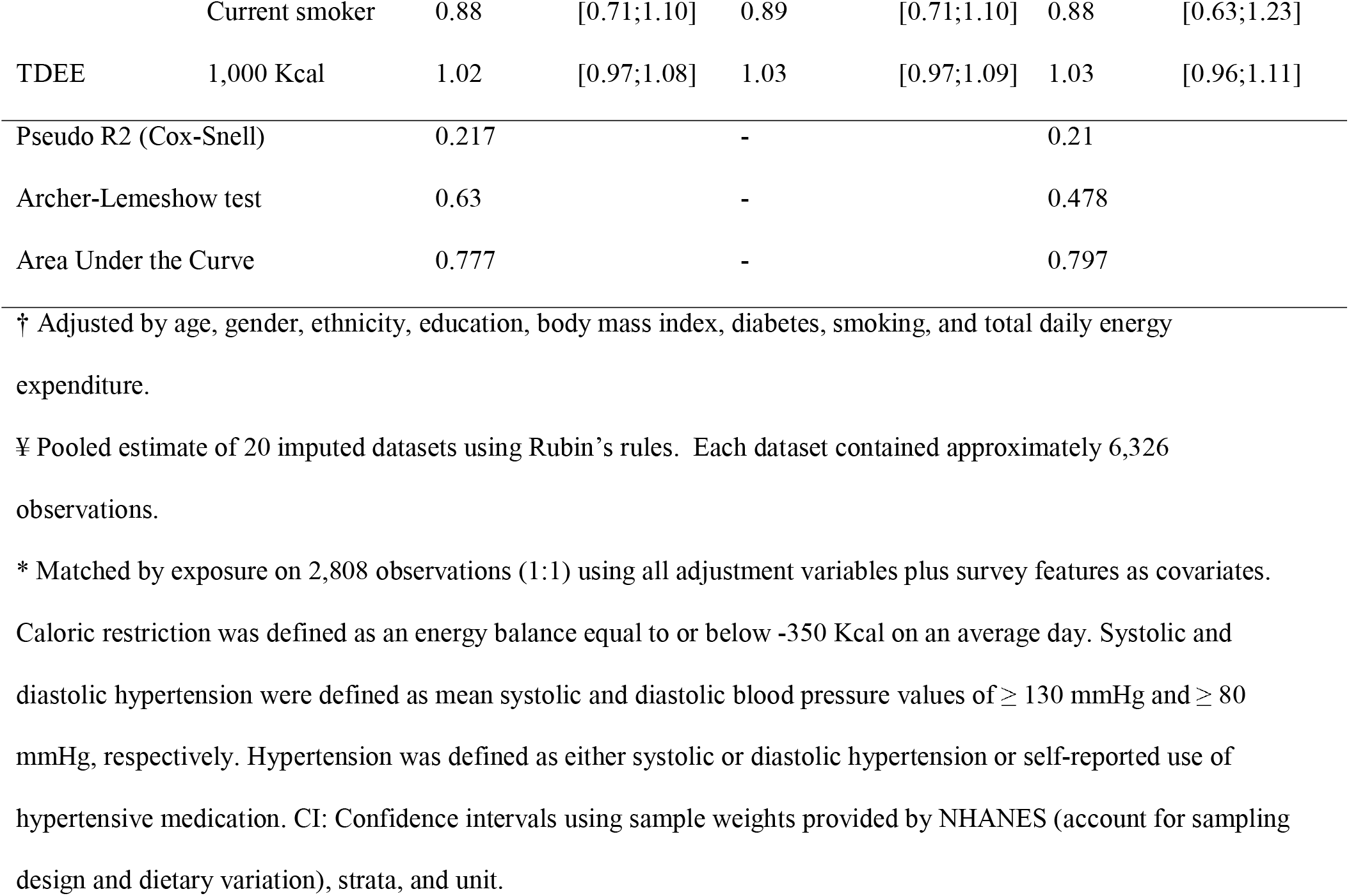
Survey-featured multivariable logistic regression model for the relationship between “high” and “low” daily sodium intake (threshold: ≥ 2.3 grams per day) and hypertension among U.S. adults in caloric restriction, aged 20-79 (National Health and Nutrition Examination Survey 2007-2018, complete-case, multiple imputations, and matched using the propensity score of the exposure.

The final propensity score for the probability of being exposed to higher amounts of sodium was estimated using age, gender, ethnicity, education, BMI (categorical), history of diabetes, history of smoking, energy expenditure, and survey features. After propensity-score matching, a total of 2,808 participants—representing a U.S. population of 24,340,468—were matched. Matching reduced the SMD to < 0.1 for all covariates, except for total daily energy expenditure (SMD = 0.27). Our outcome model corrected any remaining imbalances. The results, again, could not support an association between exposure and outcome (OR=0.99, 95% CI: 0.77; 1.27) (Table 3).

#### Sodium intake of ≥ 5.0 g/day

When the threshold for “high” sodium intake was 5 grams per day or more, the estimates did not change significantly for either the complete-case or the multiple imputation pool estimated. The lack of association, however, was accompanied by wider confidence intervals (Table 4).

We estimated the probability of being exposed to “high” sodium levels using the same model for both thresholds. A total of 926 participants—representing a U.S. population of 9,559,626—were matched by their propensity score. The SMD for all covariates was balanced (< 0.17). The binary outcome model showed no association between exposure and outcome (Table 4).

**Table 4.** Survey-featured multivariable logistic regression model for the relationship between “high” and “low” daily sodium intake (threshold: ≥ 5.0 grams per day) and hypertension among U.S. adults in caloric restriction, aged 20-79 (National Health and Nutrition Examination Survey 2007-2018), complete-case, multiple imputations, and matched using the propensity score of the exposure.

| Variable | Units | Complete |  | Multiple |  | PS- |  |
| --- | --- | --- | --- | --- | --- | --- | --- |
|  |  | case |  | Imputation |  | Matched |  |
|  |  | analysis |  | Odds |  | Odds |  |
|  |  | Ratio | CI 95% | Ratio | CI 95% | Ratio | CI 95% |
| Sodium intake | Low | Ref |  | Ref |  | Ref |  |
|  | High | 1.06† | [0.75;1.52] | 0.95†¥ | [0.67;1.35] | 1.01†* | [0.66;1.56] |
| Age | Years | 1.06 | [1.05;1.07] | 1.05 | [1.05;1.07] | 1.04 | [1.02;1.06] |
| Gender | Male | Ref |  | Ref |  | Ref |  |
|  | Female | 0.58 | [0.48;0.70] | 0.58 | [0.48;0.70] | 0.51 | [0.22;1.17] |
| Ethnicity | White | Ref |  | Ref |  | Ref |  |
|  | Latino | 0.77 | [0.62;0.94] | 0.77 | [0.63;0.95] | 0.62 | [0.34;1.13] |
|  | Black | 1.59 | [1.32;1.92] | 1.59 | [1.32;1.93] | 1.25 | [0.75;2.09] |
|  | Other | 1.23 | [0.89;1.69] | 1.23 | [0.89;1.68] | 1.07 | [0.50;2.30] |
| Education | Some school | Ref |  | Ref |  | Ref |  |
|  | High school | 0.9 | [0.68;1.19] | 0.89 | [0.68;1.18] | 0.67 | [0.35;1.30] |
|  | Higher education | 0.86 | [0.67;1.10] | 0.86 | [0.67;1.09] | 0.7 | [0.39;1.23] |
| Obese | Normal | Ref |  | Ref |  | Ref |  |
|  | Overweight | 1.39 | [1.07;1.79] | 1.37 | [1.06;1.76] | 2.19 | [1.07;4.49] |
|  | Class 1 obesity | 2.08 | [1.58;2.75] | 2.08 | [1.58;2.74] | 3.04 | [1.44;6.40] |
|  | Class 2 obesity | 2.18 | [1.59;2.98] | 2.15 | [1.58;2.94] | 3.10 | [1.40;6.91] |
|  | Class 3 obesity | 3.74 | [2.54;5.51] | 3.78 | [2.57;5.56] | 11.56 | [4.48;29.78] |
| Diabetes | No | Ref |  | Ref |  | Ref |  |
|  | Yes | 1.84 | [1.37;2.48] | 1.85 | [1.37;2.48] | 1.28 | [0.59;2.78] |
| Smoking | Never smoker | Ref |  | Ref |  | Ref |  |
|  | Past smoker | 1.05 | [0.83;1.32] | 1.05 | [0.83;1.33] | 0.85 | [0.47;1.56] |
|  | Current smoker | 0.88 | [0.71;1.10] | 0.89 | [0.72;1.10] | 0.87 | [0.47;1.61] |
| TDEE | 1,000 KCal | 1.02 | [0.96;1.08] | 1.02 | [0.96;1.08] | 0.99 | [0.91;1.07] |
| Pseudo R2 (Cox-Snell) |  | 0.216 |  | - |  | 0.15 |  |
| Archer-Lemeshow test |  | 0.89 |  | - |  | 0.47 |  |
| Area Under the Curve |  | 0.777 |  | - |  | 0.73 |  |

† Adjusted by age, gender, ethnicity, education, body mass index, diabetes, smoking, and total daily energy expenditure.
¥ Pooled estimate of 20 imputed datasets using Rubin's rules. Each dataset contained approximately 6,326 observations.
\* Matched by exposure on 926 observations (1:1) using all adjustment variables plus survey features as covariates.
Caloric restriction was defined as an energy balance equal to or below -350 Kcal on an average day. Systolic and diastolic hypertension were defined as mean systolic and diastolic blood pressure values of $\geq 130$ mmHg and $\geq 80$ mmHg, respectively. Hypertension was defined as either systolic or diastolic hypertension or self-reported use of hypertensive medication. CI: Confidence intervals using sample weights provided by NHANES (account for sampling design and dietary variation), strata, and unit.

## Discussion

### Main findings

Our analysis of a multi-year nationally representative sample of U.S. adults on caloric restriction did not detect any significant association between increased sodium consumption levels and the odds of hypertension (OR: 0.97; CI 95%: 0.90; 1.05). Our survey-featured logistic regression was adjusted by age, gender, ethnicity, education, BMI, diabetes, smoking, and total daily energy expenditure. Several sensitivity analyses yielded similar results, including those that analyzed two widely recommended sodium intake thresholds (≥2.3 and ≥5.0 grams per day). These results cement the need to target low-sodium interventions appropriately given its associated risks.^39,40^

### Contextualizing the evidence

Very few studies had been able to identify a representative sample of participants on caloric restriction, commonly identified through healthy weight loss. Our findings suggest that people on caloric restriction would see no benefit in reducing sodium in their diet to lower blood pressure. These findings apply to the general U.S. adults and other populations with similar characteristics. Our findings align with previous reports in obese adolescents, who lost their “sensitivity of blood pressure to sodium” following a 20-week intervention to lose weight (through caloric restriction).^18^

Caloric restriction lowers blood pressure levels independent of other factors; it decreases body fat and, consequentially, increases insulin sensitivity.^41^ Insulin modulates the renal absorption of sodium. As a result, people on caloric restriction excrete more sodium than those in a caloric surplus, which might explain the loss of sodium sensitivity in these patients.^42^ Such effects can be seen within hours of fasting and had been recently hypothesized as mechanisms that regulate blood pressure levels in healthy and sick individuals.^17,43,44^ While these are acute effects, other cardiometabolic benefits of caloric restriction appear to be long-lasting.^15^ We highlight the need for a tailored approach for blood pressure control in this specific subpopulation. Laxer sodium intake recommendations for these patients could help mitigate the potential low adherence to a low sodium diet and its associated adverse effects (e.g. increased risk of death).^39,40,45^ They could also reduce the complexity of dietary interventions and increase their palatability, which is paramount to improving adherence and achieving successful weight reduction.^46^

### Strengths and limitations

Our study benefited from using a population-level sample, with an adequate representation of people from diverse ethnicities, ages, and socioeconomic characteristics. Our analysis used survey features, which allowed us to generalize our findings to all the non-institutional U.S. adult population going through a healthy weight loss. We restricted our sample to participants on caloric restriction, therefore addressing the ambiguities of metabolically healthy obese patients and lean people who exhibit obese-like characteristics (dyslipidemia, altered inflammatory profile, and increased fat cell size).^47,48^ Rarely do hypertension studies account for such ambiguity. Our findings were robust under the missing at random assumption and to adjustments to the exposure to recommended intake levels by American and European current guidelines and the World Health Organization guidelines.

Our study has several limitations. We used self-reported data for energy intake, prone to measurement bias and underreporting. While our selected threshold^20^ for energy balance accounts for such underreporting, this measurement error is not systematic. We further tried to minimize bias by only including the observations marked as reliable by NHANES—providing a detailed description of each food, including the amount, additional ingredients, and eating occasions—and excluding all participants reporting a less than usual intake. NHANES interviewers also double-check to elicit forgotten foods, and the survey weights account for variation between weekends and weekdays.^49^

We also used self-reported sodium consumption instead of the 24-hour urinary collection (considered by many as the gold standard). We made this decision based on two arguments. Firstly, the Automated Multiple-Pass Method employed by NHANES provides reliable sodium and intake measures.^23^ Secondly, the hypothesized underlying mechanism (increased sodium retention) requires us to measure intake and not excretion.

Self-reported data for moderate and vigorous activity is not as reliable as objectively measured physical activity.^50^ If people reported more than 24 hours of physical and sedentary activity, NHANES analysts set those values to missing. We dealt with this first with two methods. Firstly, we took a conservative sampling (including the BMR as total energy expenditure whenever data was not available meant that participants who reported high activity levels were considered sedentary). Secondly, we used multiple imputations procedures under the assumption that these data, although not missing at random, could be predicted using the available variables.^33^ Statistically, there were no significant differences between the estimates obtained by either method. However, the direction of the effect changed with multiple imputations in both thresholds. Such a change of direction might reflect the effect of the omitted physical activity in the complete-case sample. Nevertheless, the difference was not statistically significant. Finally, our cross-sectional design did not allow us to establish a temporal relationship between exposure and outcome. It’s feasible for people to modify their diet to cope with their disease.

## Conclusion

Our findings showed that sodium intake was not associated with higher odds of hypertension among the U.S. population on caloric restriction. Our results were robust to missing data and different representations of the exposure, disputing a low-salt diet’s benefits for people who achieve weight loss and maintain it using caloric restriction. These results highlight the need to explore new population-specific strategies for sodium intake reduction, including new dietary prescription approaches that improve adherence and reduce the risk associated with deficient sodium diets.

## Data Availability

We used publicly available data from the National Health Examination and Nutrition Survey (2007-2018). R Code is available under request.

https://wwwn.cdc.gov/nchs/nhanes/

